# Heterozygous loss-of-function variants in SPTAN1 cause a novel early childhood onset distal myopathy with chronic neurogenic features

**DOI:** 10.1101/2024.09.23.24313872

**Authors:** Jonathan De Winter, Liedewei Van de Vondel, Biljana Ermanoska, Alice Monticelli, Arnaud Isapof, Enzo Cohen, Tanya Stojkovic, Peter Hackman, Mridul Johari, Johanna Palmio, Megan A. Waldrop, Alayne P. Meyer, Stefan Nicolau, Kevin M. Flanigan, Ana Töpf, Jordi Diaz-Manera, Volker Straub, Cheryl Longman, Catherine A. McWilliam, Rotem Orbach, Sumit Verma, Regina Laine, Sandra Donkervoort, Carsten G. Bonnemann, Adriana Rebelo, Stephan Züchner, Tiffany Grider, Michael E. Shy, Isabelle Maystadt, Florence Demurger, Anita Cairns, Sarah Beecroft, Chiara Folland, Willem De Ridder, Gina Ravenscroft, Gisèle Bonne, Bjarne Udd, Jonathan Baets

## Abstract

**Background:** Neurogenetic disorders caused by pathogenic variants in four genes encoding non-erythrocytic spectrins (*SPTAN1, SPTBN1, SPTBN2, SPTBN4)* range from peripheral and central nervous system involvement to complex syndromic presentations. Heterozygous pathogenic variants in *SPTAN1* are exemplary for this diversity with phenotypes spanning almost the entire spectrum.

**Methods:** Through international collaboration we identified 14 families with genetically unsolved distal weakness and unreported heterozygous *SPTAN1* loss-of-function variants including frameshift, nonsense and splice-acceptor variants. Clinical data, electrophysiology, muscle CT or MRI and muscle biopsy findings were collected and standardized. *SPTAN1* protein, mRNA expression analysis and cDNA sequencing was performed on muscle tissue from two patients.

**Results:** All 20 patients presented with early childhood onset distal weakness. The severity varied both within families and between different families. Foot abnormalities ranged from hammer toes and pes cavus to distal arthrogryposis. Electrophysiology showed mixed myogenic and neurogenic features. Muscle MRI or CT in 10 patients showed fatty infiltration of the distal lower limb anterior compartment and/or selective involvement of the extensor hallucis longus muscle. Muscle biopsy revealed myopathic changes with mild dystrophic and chronic neurogenic changes in 7 patients. Finally, we provide proof for nonsense mediated decay in tissues derived from two patients.

**Conclusions:** We provide evidence for the association of *SPTAN1* loss-of-function variants with childhood onset distal myopathy in 14 families. This finding extends the phenotypic spectrum of *SPTAN1* loss-of-function variants ranging from intellectual disability to distal weakness with a predominant myogenic cause.

**KEY MESSAGES:**

⋄ *SPTAN1* loss-of-function variants, including frameshift, nonsense and splice site variants cause a novel childhood onset distal weakness syndrome with primarily skeletal muscle involvement.
⋄ Hereditary motor neuropathies and distal myopathic disorders present a well-known diagnostic challenge as they demonstrate substantial clinical and genetic overlap. The emergence of *SPTAN1* loss-of-function variants serves as a noteworthy example, highlighting a growing convergence in the spectrum of genotypes linked to both hereditary motor neuropathies and distal myopathies.

## INTRODUCTION

Heterozygous variants in *SPTAN1* can cause a wide spectrum of inherited neurological disorders and are a prime example of the daily challenges both bedside and laboratory practices face in the complex domain of rare inherited neurological disorders. Further advancements in our understanding of neurogenetic disorders relies heavily on international collaborations. Herein, the emphasis lies in solving the genetically unsolved patients by connecting patient cohorts and research groups. Through these international efforts the pool of disease-associated genes is consistently growing, with for example over 300 genes identified for inherited myopathies. These genetic insights revealed several pathomechanistic pathways and key protein families that are often shared amongst seemingly different neurological disorders such as hereditary spastic paraplegia (HSP), cerebellar ataxia (SCA) and inherited peripheral neuropathy (IPN).^1^ Pathogenic variants in spectrin genes, such as *SPTAN1*, present a well-established group of disorders ranging from central to peripheral nervous system involvement, also referred to as ‘spectrinopathies’.^2^

The spectrin proteins are integral part of the submembranous cytoskeleton in metazoan cells.^3^ Alpha-1-spectrin (*SPTA1*) and beta-1-spectrin (*SPTB1*) are expressed in red blood cells and both are associated with inherited red blood cell disorders, namely hereditary elliptocytosis and spherocytosis.^4^ Alpha-2-spectrin (*SPTAN1*) and four different beta-spectrins (*SPTBN1, SPTBN2, SPTBN4, SPTBN5)* show widespread expression, yet are notably enriched in neuronal tissues.^2^ Recent advances in super-resolution microscopy helped to identify the axonal submembranous cytoskeleton organized as ~190 nm periodic repeats of spectrin and actin rings, providing a mechanical scaffold and accurate subcellular positioning of various crucial client proteins or protein complexes such as ankyrins, ion channels and transmembrane adhesion molecules.^5,6^ The basic axonal spectrin units are heterodimers consisting of SPTAN1 and one of the neuronal beta-spectrins that self-associate into heterotetramers with “tails” that crosslink actin filaments and enable the periodic organization (Figure 1). Thus, alpha-2-spectrin is a crucial organizer of the axonal cytoskeleton.

**Figure 1:**
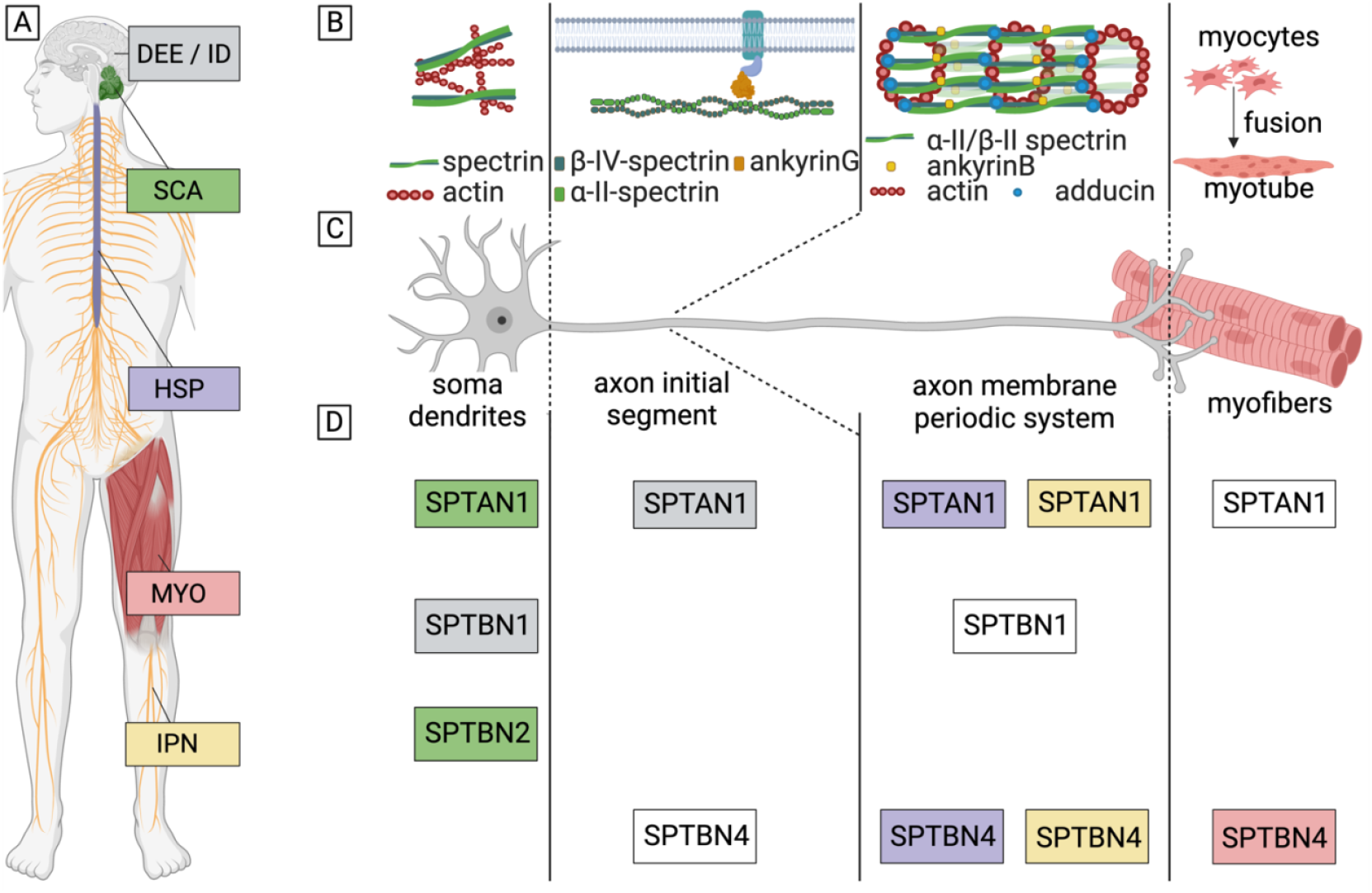
Neurospectrinopathy spectrum and spectrin protein functions. Graphical representation of the neurospectrinopathy genotype and phenotype spectrum (A and D), spectrin protein functions (B) and spectrin subcellular localization (C and D). A: spectrin phenotypic spectrum ranges from developmental and epileptic encephalopathy (DEE), intellectual disability (ID), to spinocerebellar ataxia (SCA), hereditary spastic paraplegia (HSP), myopathy (MYO) and inherited peripheral neuropathy (IPN). B: function of the alpha-2-spectrin depends on the subcellular localization (C). In the soma and dendrites spectrin complexes interact with actin to form a mesh-like structure. At the axon initial segment spectrin complexes display an essential scaffolding function, providing the correct localization of ankyrins and ion channels (not displayed in the figure). In the axon spectrin interacts with actin rings, ankyrins and adducin to form the membrane periodic system. At the level of the myofibers spectrin is shown to be involved in the process of myofiber regeneration. D: subcellular localization and phenotypic association of the different spectrins: *SPTAN1, SPTBN1, SPTBN2* and *SPTBN4*. The colored boxes represent the different phenotypes: grey: DEE/ID, green: SCA, purple: HSP, red: MYO, yellow: IPN, white box: no disease association so far which can be linked to that specific subcellular region. The function and localization of *SPTBN5* has not been uncovered in the current literature.

Interestingly, pathogenic variants in both alpha-2-spectrin and the different beta-spectrins have been reported to cause neurological disorders ranging from epilepsy syndromes, HSP, SCA, intellectual disability (ID) to IPN, highlighting their important role in maintaining neuronal health (Figure 1). A total of 60 different heterozygous *SPTAN1* pathogenic variants are reported to cause early childhood to juvenile onset of neurological dysfunction (Figure 2). Although establishing clear genotype-phenotype correlations remains challenging, certain clinical presentations seem to associate with specific types of *SPTAN1* variants and hint towards a possible classification of *SPTAN1-*related disorders into three primary groups. One group could consist of severe developmental and epileptic encephalopathy syndromes and is mainly associated with missense and small in-frame indel variants occurring in the last two spectrin repeats. These pathogenic variants likely impair alpha-2-spectrin heterodimerization with one of the beta-spectrins and were shown to cause protein aggregation for the most recurrent variant, namely p.(Asp2303_Leu2305dup).^7^ The second group could include patients with less severe forms of epilepsy, ID and pure or complicated forms of HSP and SCA. Herein, we reported on missense mutations and in-frame deletions likely causing a destabilization of interlinking spectrin helices.^8^ This destabilization could be reflected by the protein aggregation found in patient-derived fibroblasts for the most recurrent *SPTAN1* variant associated with HSP, namely p.(Arg19Trp).^9^ Finally, *SPTAN1* nonsense and frameshift variants were reported to cause hereditary motor neuropathy (HMN) or ID with varying intra- and interfamilial severity *via* haploinsufficiency caused by the nonsense-mediated decay (NMD) pathway.^9,10^ It is worthwhile to mention that these three groups of *SPTAN1-*related disorders span the entire spectrinopathy spectrum.

**Figure 2:**
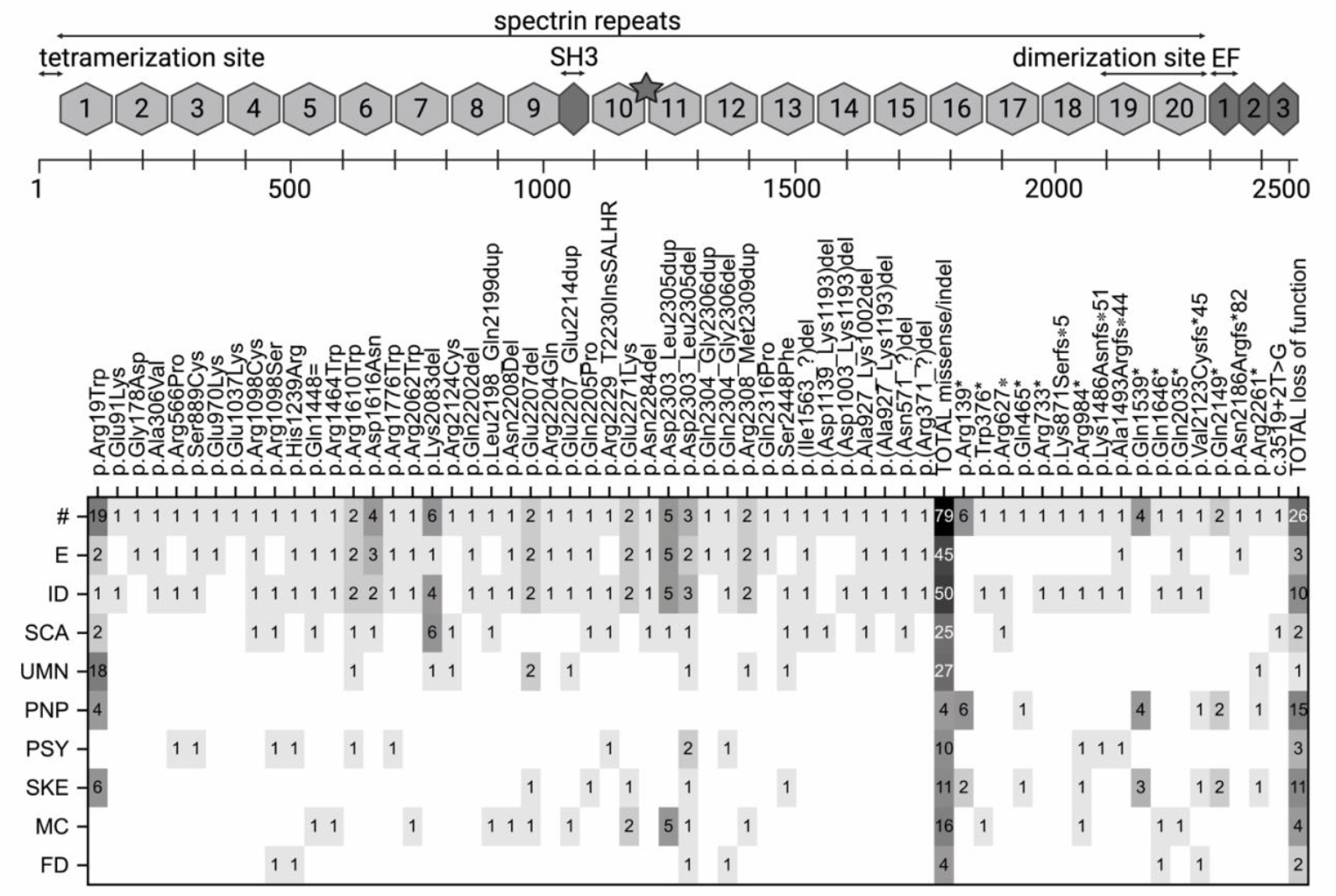
*SPTAN1* reported variants and phenotypic associations. Top part of the figure shows a schematic representation of the alpha-2-spectrin protein (grey star indicates calmodulin binding site and calpain cleavage domain). Bottom part of the figure shows a heatmap including all 60 different *SPTAN1* variants reported and number of patients (#) to cause epilepsy and intellectual disability, cerebellar ataxia, spastic paraplegia and peripheral neuropathy. Within the group of *SPTAN1* missense and indel variants the p.Arg19Trp, p.Lys2083del and p.Asp2303_Leu2305dup variants are highly recurrent and are associated with hereditary spastic paraplagia, spinocerebellar ataxia and developmental and epileptic encephalopathy respectively. In total 17 different loss-of-function variants (nonsense, frameshift and splice variant) were mainly reported with a varying severity of intellectual disability and/or distal weakness syndrome. Abbreviations: #: number of patients reported; E: epilepsy; ID: intellectual disability; SCA: spinocerebellar ataxia; UMN: upper motor neuron involvement; PNP: peripheral neuropathy; PSY: psychiatric symptoms and signs; SKE: skeletal abnormalities including foot, hand abnormalities and scoliosis; MC: microcephaly; FD: facial dysmorphism.

In this study, we identified 14 families with novel heterozygous *SPTAN1* loss-of-function variants presenting with an early childhood onset distal muscle weakness syndrome characterized by myopathic abnormalities and to a lesser extent chronic neurogenic findings on muscle biopsy. This expansion of the *SPTAN1*-associated disease spectrum demonstrates that different tissues could be differentially affected by heterozygous loss-of-function variants in this essential, ubiquitous cytoskeletal protein.

## METHODS

### Patient recruitment and expert consensus on phenotype

Through collaboration within the Solve-RD EU-Horizon 2020 project (https://solve-rd.eu/), Genesis platform (https://tgp-foundation.org) and by expert referrals we identified 14 families presenting with distal weakness that carry heterozygous *SPTAN1* loss-of-function variants. All participating patients and/or their legal representatives gave written informed consent locally and received clinical evaluations as part of standard care in centers specialized in neurology and neuromuscular disorders. Clinical data were collected by a standardized document and subsequently discussed by the different referring centers to reach consensus on the interpretation of clinical features, electrophysiology, muscle MRI (axial T1-weighted images) or CT and muscle biopsy if available. When available muscle MR images were evaluated to calculate the modified Mercuri score. This score is a subjective quantification of fatty replacement in skeletal muscle ranging from 0 to 4 based on axial T1-weighted images.

### Genetic studies

Whole exome sequencing was performed by conventional next-generation sequencing technologies using locally applied protocols and shared by the Solve-RD project through Genome-Phenome Analysis Platform (GPAP) or on the Genesis platform. Filtering for heterozygous loss-of-function variants was performed using standard quality filtering settings. Where possible, Sanger sequencing was performed to extend segregation analysis. All *SPTAN1* variants were mapped to transcript NM_001130438.2. Diagnostic work-up showed that none of the patients carried a likely pathogenic or pathogenic variant in other genes known to cause inherited distal weakness syndromes, including both HMN and distal myopathy (dMYO) related genetic etiologies.

### Experimental studies

In two families, muscle tissue of the affected probands (B:II:1 and N:II:1) was collected for experimental analysis. *SPTAN1* protein, mRNA expression and cDNA sequencing were performed on muscle tissue from B:II:1. Western blotting was performed on homogenized muscle tissue lysed in RIPA buffer. A 4-12% gradient gel was used, followed by transfer and blocking. Imaging was performed using mouse anti-SPTAN1 antibody (1:1000) (Abcam ab11755) and GAPDH (Genetex GTX100118 1:10 000) as a loading control. RNA was extracted from the muscle tissue using the Universal RNA kit (Roboklon) and subsequently cDNA was generated for qPCR testing using the High-Capacity cDNA Reverse Transcription Kit (ThermoFisher) both according to the manufacturer’s protocol. RT-qPCR was performed using the SYBR Green PCR Master Mix (Fisher Scientific), *SPTAN1* qPCR primers (F: CTGAAGGTCTCATGGCAGAGGA,R:CACGGTGTGAACCATCAGACGA) and *GAPDH* qPCR primers (F: TGCACCACCAACTGCTTAGC, R: GGCATGGACTGTGGTCATGAG). Sanger sequencing around the mutation of interest in the generated cDNA was performed to investigate targeting of the mutant mRNA by NMD, using the following *SPTAN1* primers (F: CGACACTCTTGCCGCCATCC, R: GCAGCCCAGCGTCAAAAGTT). Finally, RNA was extracted from muscle tissue from N:II:1 for *SPTAN1* RNA sequencing analysis using the DROP pipeline (supplementary file).^12^ Sashimi plots were created to interpret the effect of the *de novo* splice acceptor variant in N:II:1 (Figure 6).

## RESULTS

### Genetic evidence

All 20 affected patients carried heterozygous *SPTAN1* loss-of-function variants, more specifically nine frameshift variants, four nonsense variants and one splice-acceptor variant (Figure 3). Five families (A, B, C, L and M) showed autosomal dominant mode of inheritance, while in nine patients the variant was shown to be *de novo*, including 2 pairs of monozygotic twins (J, K). In families E and G further segregation analysis in the unaffected parents was not possible and therefore the inheritance mode remains unknown without proof of a suspected *de novo* inheritance. In total, seven European families (A, B, C, F, G, L and M), six American (D, E, H, I, J, K) families, and one Australian family (N) were included. All variants were absent in the healthy control population in GnomAD version 4 (https://gnomad.broadinstitute.org) and were situated outside the N-terminal domain or last exon of *SPTAN1* suggesting the susceptibility of the mutated mRNA product to NMD. Moreover, *SPTAN1* has a high predicted loss-of-function intolerance score (pLI) of 1 (with pLI > 0.9 is already considered extremely intolerant for loss-of-function variants), supporting further the putative pathogenicity of the heterozygous *SPTAN1* loss-of-function mutations identified in this study.

**Figure 3:**
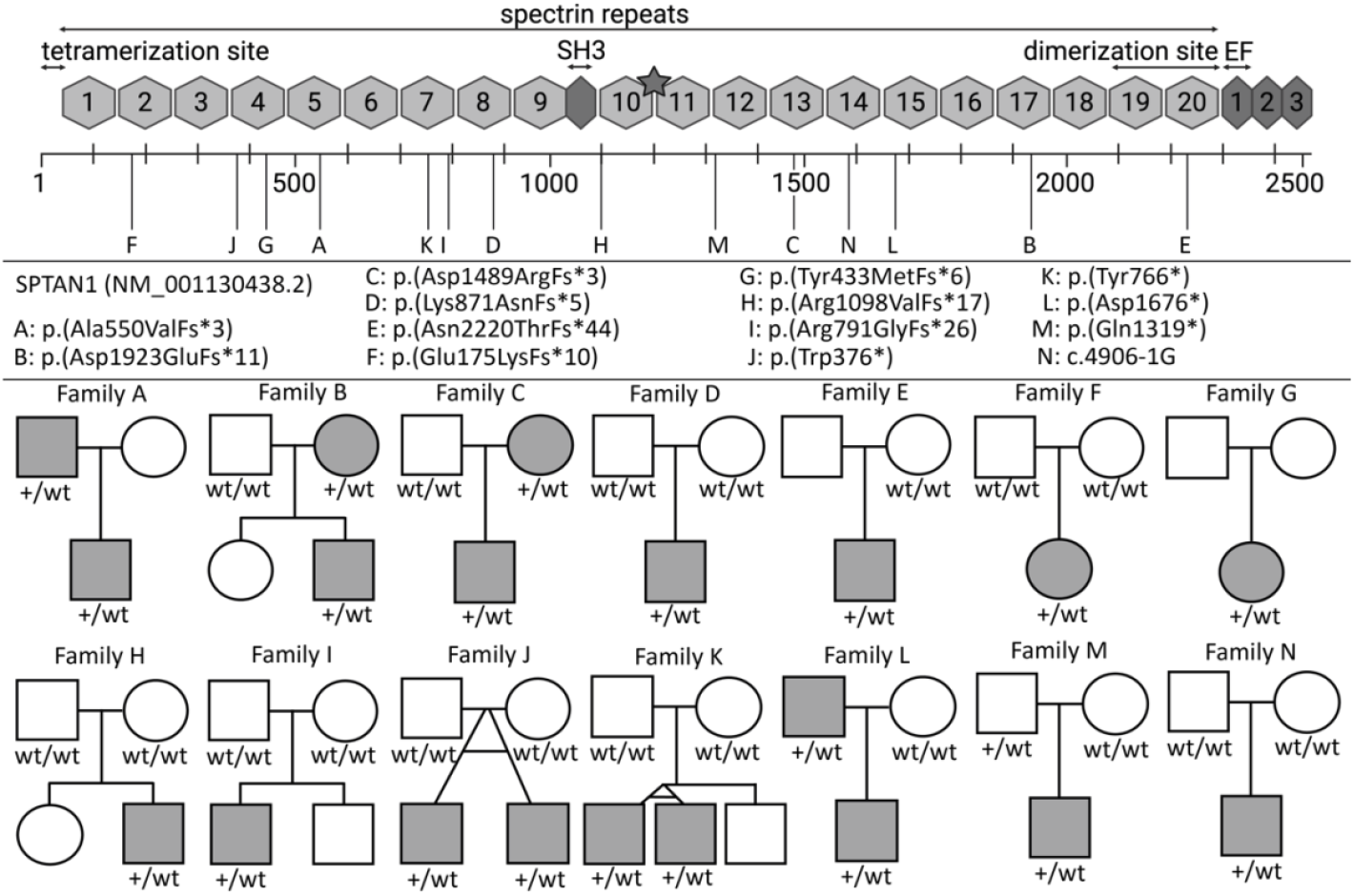
Pedigrees of families with *SPTAN1* loss-of-function variants. Pedigrees of families A – N with their respective *SPTAN1* variants, including *SPTAN1* frameshift variants (A-I), nonsense variants (J-M) and splice acceptor variant (N). Affected individuals are indicated in grey, unaffected individuals in white. Abbreviations: wt: wild type allele; + mutated allele.

### Clinical findings

We describe 20 patients belonging to 14 unrelated families, all presenting with early childhood onset of gait and foot abnormalities. In general, patients showed stable or very slowly progressive (9/20) distal muscle weakness involving the foot (19/20) and toe extensors (15/20). Of note two patients (B:I:1 and L:I:1) did not report any gait disturbances but were noted to have a left-sided selective weakness of the extensor hallucis longus muscle (B:I:1) or mild distal muscular amyotrophy and subtle weakness of foot and big toe extensors bilaterally (L:I:1). Patient M:I:1 reported no signs of weakness or foot abnormalities but was not formally clinically investigated and therefore could indicate non-penetrance. Proximal muscle involvement was relatively uncommon and included mild hip flexion weakness (4/20), bulbar muscle involvement (4/20) and neck flexion weakness (2/20). None of the patients showed signs of cardiac involvement. Foot abnormalities were present in ten patients and ranged from pes cavus (5/20) or pes planovalgus (1/20) to distal arthrogryposis (4/20). Scoliosis was observed in two patients (A:I:1 and B:II:1). Microcephaly was noted in patient A:I:1 and also in patient M:II:1. None of the patients showed hearing or vision loss or features of pyramidal tract or cerebellar involvement. Clinical details are summarized in table 1.

**Table 1:**
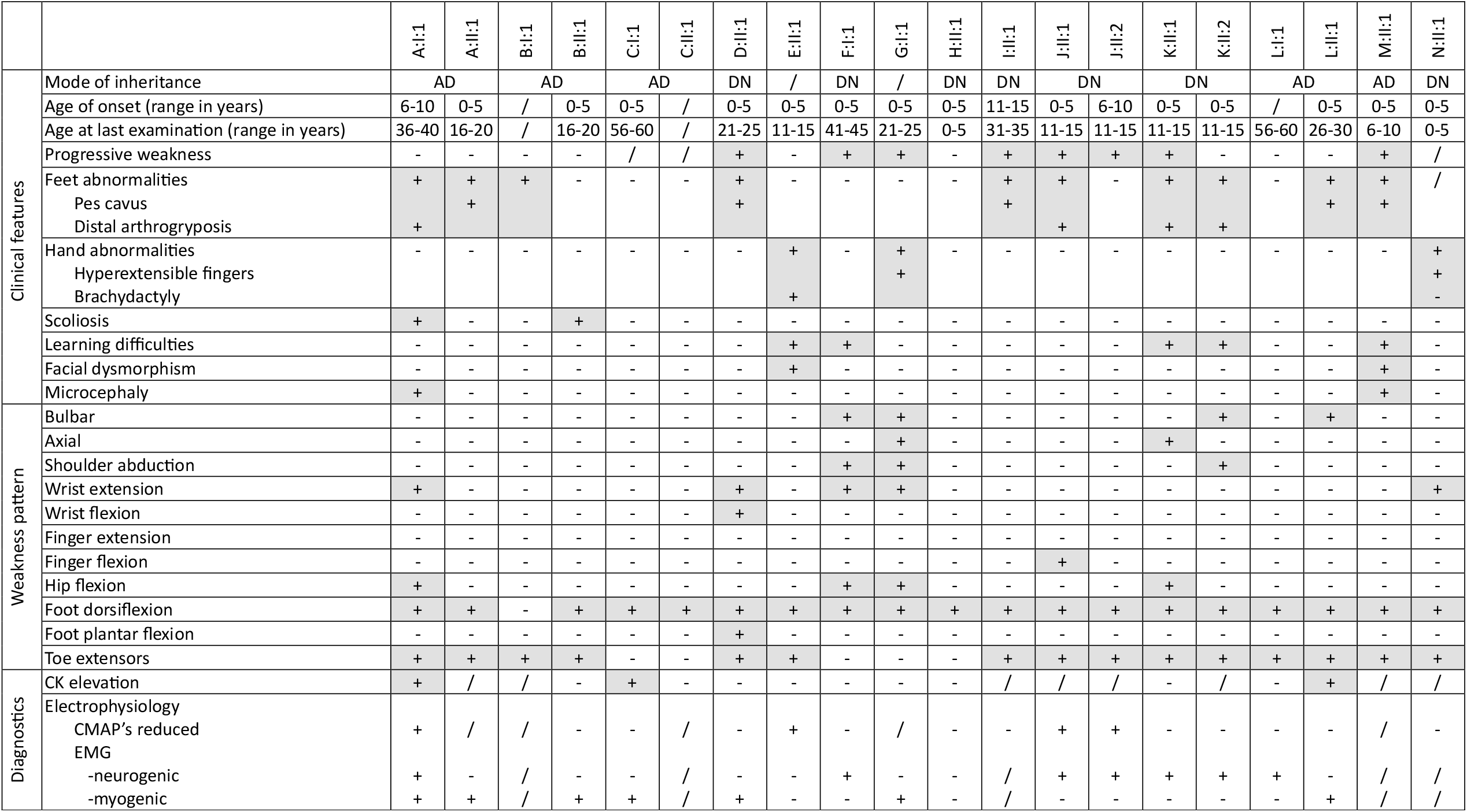
Overview of clinical features of patients carrying *SPTAN1* loss-of-function variants. Abbreviations: +: present; −: absent; /: no data available; AD: autosomal dominant; DN: *de novo*.

### Diagnostic findings

When available, patients showed normal to mildly elevated serum creatinine kinase (CK) levels (1.5-2 times upper limit). For most patients, nerve conduction studies were in line with preserved motor nerve amplitudes (11/14), while only in four patients reduced amplitudes were detected. Electromyography showed varying neurogenic, myogenic or mixed patterns. Ultrasound examination of the tibial anterior muscle in patient K:II:1 showed a Heckmatt grade 2 granular increased echogenicity indicating the presence of increased fibrous tissue and fat infiltration in the examined muscle. Muscle MRI was obtained in nine out of 20 patients, with muscle fatty replacement predominantly affecting muscles of the distal anterior leg compartment (Figure 4). Selective extensor hallucis longus muscle involvement was shown as a so-called ‘slit sign’ in six patients. This radiological sign can be seen as an empty space between the tibialis anterior and the extensor digitorum longus muscle. Importantly, the presence of the slit sign in patient B:I:1 correlated with the unilateral hanging toe sign encountered during clinical examination. Proximal muscle involvement was noted in patients A:II:1 and G:I:1. In general affected muscles did not show reticular involvement or so-called muscle islands or pop-corn appearance such as in inherited motor neuro(no)pathies.^13^ Muscle biopsy was performed in 7/20 patients, in which myopathic changes were present in all samples, including an increase in fiber size variation (7/7), marked muscle fiber hypertrophy in at least two patients, increase in internal nuclei (4/7), fiber splitting (6/7), increase in endomysial fibrosis (5/7) and some regenerating fibers (3/7). The identification of fiber type grouping (6/7) together with the presence of some angulated atrophic fibers (6/7) in some patients implied a concurrent chronic denervation process, though its presence is comparatively less pronounced than the observed myopathic changes. There were no signs of active inflammation, target fibers, myonecrosis or vacuolar changes. Of note, six families were initially considered as HMN (A, G, I, J, L and N), four of which were reclassified as distal myopathy based on the muscle biopsy findings (Figure 5).

**Figure 4:**
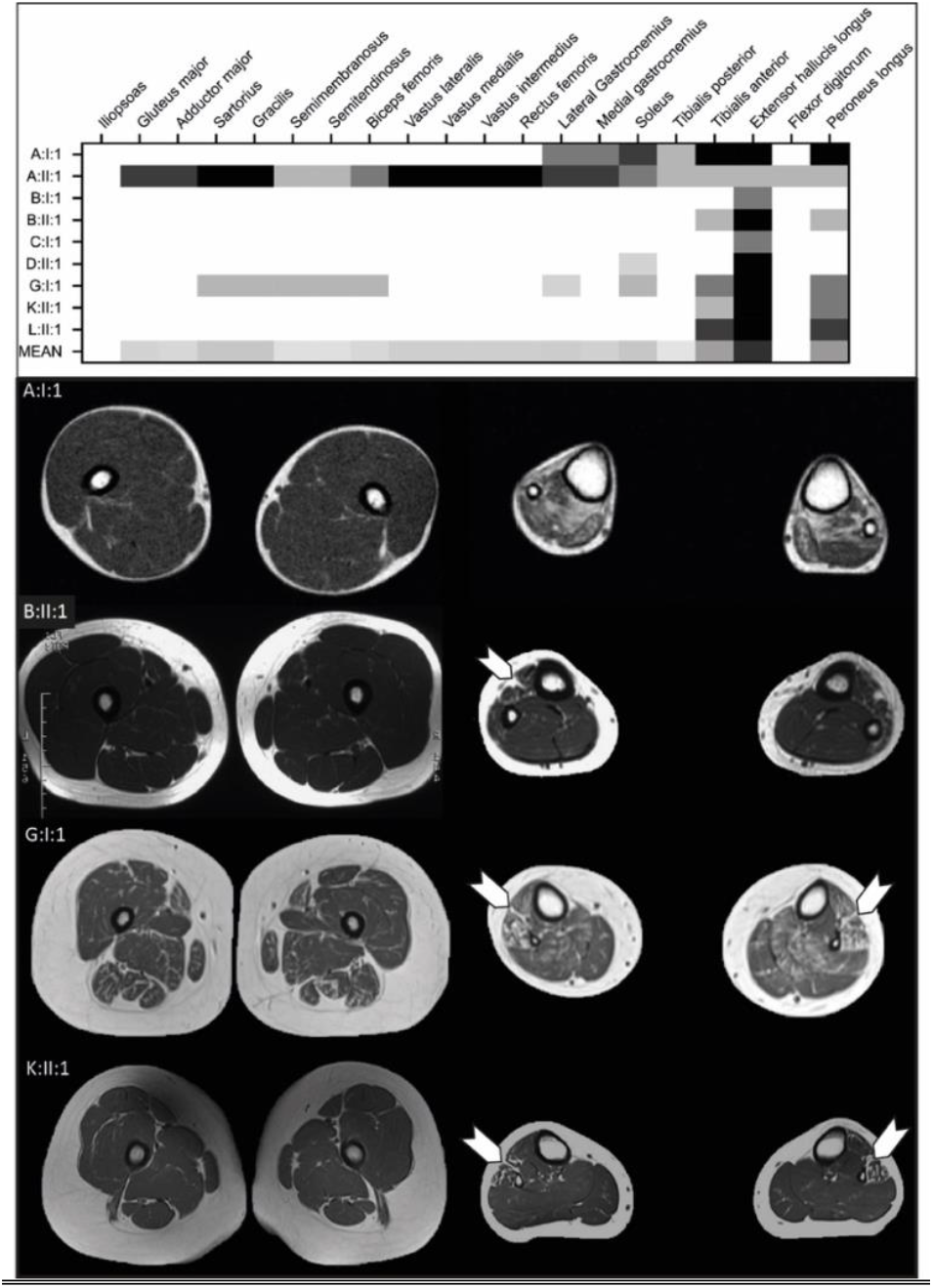
Heatmap of the lower limb modified Mercuri score and representative T1 weighted muscle MRI images. In 9 patients carrying different *SPTAN1* loss-of-function variants modified Mercuri score was calculated representing fatty degeneration of lower limb muscles. Most patients showed predominant involvement of the anterior compartment of the distal lower limbs, including most frequently tibialis anterior, peroneus longus and the extensor hallucis longus (EHL) muscle. The slit sign is indicated by the white arrowheads and implies the absence of the EHL muscle. Less frequently the posterior compartment of the proximal lower limbs was involved (A:II:1 and G:II:1).

**Figure 5:**
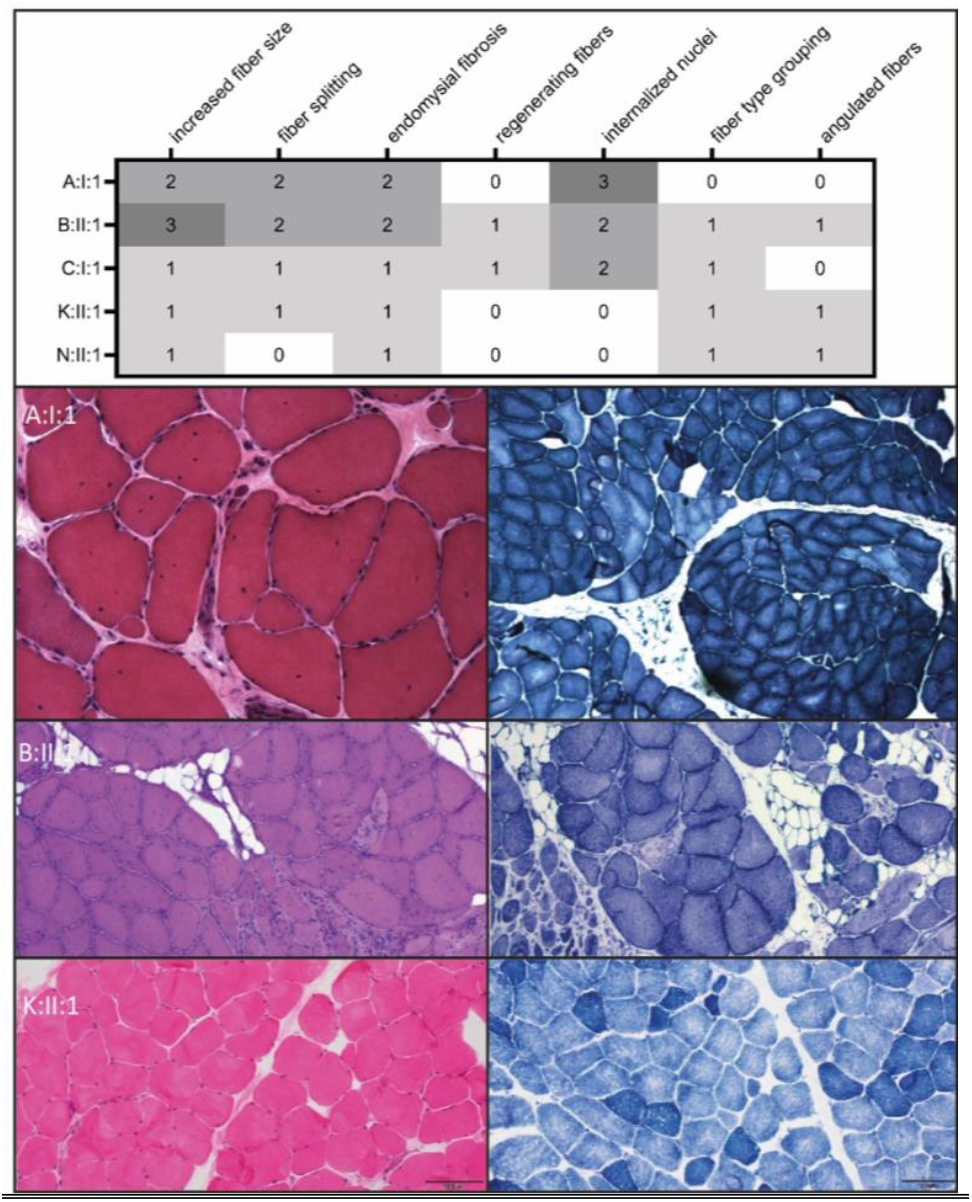
Heatmap summarizing muscle biopsy findings in patients with *SPTAN1* loss-of-function variants combined with representative microscopy images. The heatmap indicates the presence of myopathic changes in 5 patients from whom muscle biopsy images were available to evaluate. Mild chronic denervating features (fiber type grouping, angulated atrophic fibers) are present to a lesser extent. Relative severity scores were given ranging from 0 to 3 (0: absent; 1: discrete; 2: evident; 3: profound). Lower panels include representative microscopy images from patients A:I:1 (deltoid muscle), B:II:1 (tibialis anterior muscle), K:II:1 (tibialis anterio muscle). Left panels show the hematoxylin and eosin (H&E) stainings, the right panels include oxidative stainings (NADH-TR).

**Figure 6:**
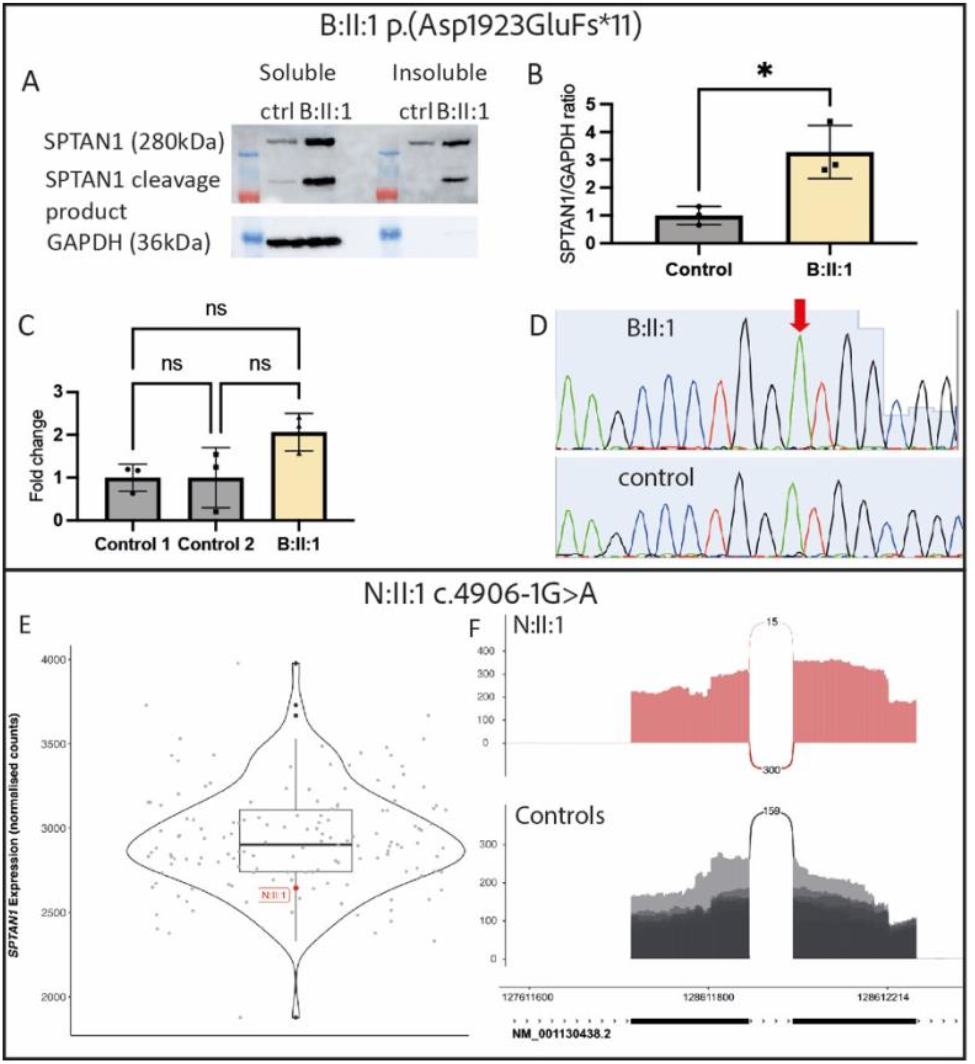
Experimental assessment on muscle tissue from patient B:II:1 and N:II:1. Upper and lower row includes experimental results from patient B:II:1 and N:II:1, respectively. Muscle tissue from both patients were derived from tibial anterior muscle. A: Muscle protein expression on western blot probing for SPTAN1 and GAPDH (loading control) (cropped images), including soluble and insoluble fractions. B: quantification and statistical analysis (unpaired t-test) of SPTAN1 and GAPDH protein concentrations. C: quantification and statistical analysis (one-way ANOVA) of *SPTAN1* gene expression levels normalized to *GAPDH*. D: cDNA Sanger sequence traces showing only the presence of the wild type allele indicative for the presence of NMD. E: *SPTAN1* RNA expression of N:II:1 (red dot) compared to 149 skeletal muscle RNA-seq from rare muscle disease patients and unaffected controls. F: Sashimi plots show the use of an alternate acceptor site in N:II:1 (red), compared to six unaffected controls (grey). The alternate acceptor is at position g.128612109 and is supported by 15 reads. Abbreviations: ctrl, control. Levels of statistical significance p-value: non-significant (ns) ≥ 0.05, significant (*) 0.01-0.05, very significant (**) 0.001-0.01, extremely significant (***) 0.0001-0.001.

In conclusion, the clinical presentation combined with the radiological and histopathological findings point toward a novel congenital distal myopathy with dystrophic changes and discrete chronic neurogenic features.

### Experimental evidence

Previously, we identified *SPTAN1* nonsense variants to cause HMN in three independent families and showed evidence for haploinsufficiency by western blot analysis, mRNA PCR and Sanger sequencing, and cycloheximide treatment of patient-derived lymphoblasts.^10^ In this study we analyzed available muscle tissue from two unrelated patients carrying a *SPTAN1* frameshift variant (B:II:1) and a splice acceptor variant (N:II:1). We found significantly (p-value = 0.0173) elevated alpha-2-spectrin protein levels in patient B:II:1 compared to healthy controls, without evidence of both full length or truncated alpha-2-spectrin accumulation in the insoluble fraction. In contrast, we observed a non-significant increase (p-value = 0.069) in total *SPTAN1* mRNA expression in patient B:II:1.

Finally, cDNA sequencing showed absence of the mutant allele in the affected muscle tissue from patient B:II:1 confirming the frameshift (c.5767_5768insA, p.Asp1923GluFs*11) variant triggered the NMD mechanism (Figure 6). Finally, we performed RNA sequencing on skeletal muscle RNA derived from patient N:II:1 carrying a *de novo* c.4906-1G>A for further experimental analysis. Sashimi plots showed the use of an alternate acceptor site at position g.128612109 which was supported by 15 reads. The alternative splicing removes a single guanine nucleotide, resulting in a frameshift variant (p.Ala1636Profs*5). Analysis using OUTRIDER (as part of DROP) did not identify any expression outliers. Expression of *SPTAN1* was normal compared to unaffected controls and patients with unrelated neuromuscular disorders (Figure 6).

## DISCUSSION

We describe 20 patients from 14 unrelated families carrying novel *SPTAN1* loss-of-function variants with an early childhood onset distal weakness phenotype. By deep phenotyping including whole body muscle MRI and muscle biopsy findings, we provide support for a novel distal weakness syndrome that is a distal myopathy subtype characterized by mild dystrophic findings often mixed with varying degrees of chronic neurogenic changes. In two patients we show evidence of NMD in patient muscle tissue, yet with normal to increased protein levels and normal mRNA expression.

Initially, we reported *SPTAN1* nonsense variants to cause HMN in three unrelated families, a finding which was later replicated in two independent reports.^10,14,15^ The cohort included in the current study exhibits several similarities to these previously reported patients but presents multiple arguments supporting a diagnosis of a congenital distal myopathy rather than HMN. First, the clinical presentation of distal weakness, along with variable generalized muscle weakness including mild facial muscle involvement, and a very slow or absent disease progression, strongly indicates a primary myopathic origin. Notably, like previously reported patients, the severity of muscle weakness is highly variable both within and between families. For example, patient B:I:1 exhibits only a left-sided drop toe sign, whereas patients F:I:1 and G:I:1 display generalized muscle weakness. Additionally, the severity of foot abnormalities ranges from hammer toes to distal arthrogryposis. Further technical evaluation of our cohort showed additional proof for a novel type of distal myopathy. Herein, CK-levels were normal to mildly elevated (1,5 – 2x upper limit), while electrophysiological examination predominantly pointed towards myogenic with or without neurogenic abnormalities. Muscle MRI or CT showed involvement of the distal anterior compartment of the legs in seven patients, with relative sparing of the posterior compartment. Of note, selective fatty replacement or absence of the extensor hallucis longus muscle was seen in B:I:1, C:I:1, D:II:1, G:I:1 and L:II:1. Coined as the ‘slit sign’, this is a non-specific finding but can indicate a primary myopathic process since it can be seen as the clinical ‘hanging big toe sign’ in other distal myopathies such as those caused by *MYH7, NEB* and *MYPN*.^16^ Also, the EHL was shown to be least affected on MRI in a cohort of 84 patients with HMN, rendering a primary myopathic origin in our cohort more likely. Other neurogenic features such a reticular pattern or muscle islands were less clearly present in our cohort.^17^

Muscle biopsies from seven patients provided additional evidence that *SPTAN1* loss-of-function variants cause a distal myopathy by revealing a marked increase in internalized nuclei, muscle fiber splitting, regenerating fibers, and endomysial fibrosis. Together with the presence of marked muscle fiber hypertrophy these are pathological features reminiscent of congenital myopathies.^18^ Chronic neurogenic changes, such as fiber type grouping and angulated atrophic fibers, were present to a lesser extent, suggesting a concomitant mild chronic denervation process in some patients. In the literature, several reports touched upon this topic when – sometimes unexpectedly – myopathic findings are found on muscle biopsy in patients considered to be affected with HMN, such as seen in the heat-shock proteins (*HSPB1, HSPB3, HSPB8, DNAJB2*) and other genes including *BICD2, KIF5A, MYH14, SOD1, HINT1* and *VWA1*. It is debated whether the myopathic changes can be considered as a secondary phenomenon. Often reports refer to a historic study of muscle histopathological examinations in post-poliomyelitis patients with residual muscle weakness. In this study a surprisingly high degree of myopathic alterations was found which, according to the authors, could not otherwise be explained as secondary.^19^ However, it remains uncertain in literature why certain HMN related genes associate with myopathic changes and why others do not. Altogether, the consistent clinical presentation of slowly to non-progressive muscle weakness, predominantly affecting distal muscles and, in some cases, more generalized weakness, combined with muscle MRI findings and myopathic alterations observed on muscle biopsy, indicates that this distal weakness syndrome is a novel type of early-onset distal myopathy with signs of neuropathic overlap. This also indicates that myopathic changes could be missed in patients presenting with a distal weakness syndrome, a neurogenic EMG and no muscle biopsy available. Therefore, it is reasonable to think that the previous patients we reported with HMN, could in fact have muscle involvement as well.^10^ These patients were however not available for muscle MRI or biopsy. In addition to the reported HMN phenotype caused by *SPTAN1* nonsense variants, patients carrying heterozygous *SPTAN1* loss-of-function variants are reported to present with intellectual disability, of which, two patients were reported to have distal weakness. In our cohort, five patients show signs of very mild cognitive involvement, namely dyscalculia in patient F:I:1 and delayed language acquisition in patients E:II:1, K:II:1, K:II:2 and M:II:1. Thus from a clinical point of view, all patients showed an early childhood onset of weakness with little to no disease progression and some showing arthrogryposis and/or mild intellectual disability. These observations suggest the manifestation of a developmental distal myopathy rather than a degenerative disorder.

We provide strong genetic evidence for the pathogenicity of *SPTAN1* loss-of-function variants by including in total seven families with a *de novo* mode of inheritance, including two sets of homozygotic twins. *SPTAN1* haploinsufficiency is a well-established mutational mechanism as is reported in the initial HMN cohort.^10^ In this cohort we encountered *SPTAN1* mRNA levels in muscle tissue in patient N:II:1 comparable to *SPTAN1* mRNA levels of other affected and unaffected controls. Moreover, we found evidence of NMD by cDNA sequencing, but unexpected elevated *SPTAN1* mRNA and protein levels in patient B:II:1 compared to healthy controls. Interestingly, patients B:II:1 and his mother B:I:1 are clearly less affected in terms of clinical severity when compared to N:II:1. Altogether, these results indicate that the nonsense and frameshift variants encountered in these families are susceptible to NMD and the loss of these alleles could be transcriptionally compensated. This compensation mechanism is a known mechanism based on RNA decay as a trigger to increase expressivity of the wild-type allele.^20^

We hypothesize that loss-of-function variants in *SPTAN1* causing this novel distal myopathy could be considered a developmental syndrome, where a bottleneck in spectrin availability during early development is the underlying cause. Spectrins are crucial during myoblast fusion due to their mechanoresponsive capabilities.^21^ Also, during embryonic skeletal muscle development spectrins were shown to associate with laminin-alpha-5 and CD239 thereby bridging the muscle fiber basement membrane with the intracellular cytoskeleton.^22^ Therefore, the combination of relatively low alpha-2-spectrin expression in muscle and the loss-of-function variants in our cohort likely leads to an insufficient supply of spectrin during high cellular demand, such as during the development of the musculoskeletal system. Also, differences in NMD efficiency together with the intrinsic demands of different cell types and capability to transcriptionally compensate could explain the observed tissue-specific phenotype and possibly even the intra- and inter-familial variability in severity of the phenotype.^23^ Further experimental research is needed to validate this hypothesis and could provide valuable insights towards other congenital or early-onset distal myopathies.

*SPTAN1* loss-of-function variants causing a developmental distal myopathy phenotype adds further complexity to the phenotypic spectrum of *SPTAN1*-related disorders. However, within the broader spectrin protein superfamily comparable phenotypes are associated with a loss-of-function mutational mechanism. Biallelic loss-of-function variants in *SPTBN4* cause a mixed neuropathy-myopathy phenotype with intellectual disability, albeit more severe than we encountered in our cohort. Heterozygous loss-of-function variants in *SPTBN1* are associated with a neurodevelopmental syndrome with variable features of epilepsy, facial dysmorphisms and hypotonia. It is worthwhile to mention that muscle MRI or biopsy was not performed in these patients nor implemented in the mouse model.^24^ Finally, a homozygous *SPTBN5* nonsense variant was identified in a patient affected with intellectual disability, seizures, facial dysmorphic features and clinodactyly. However, the report did not include muscle strength evaluation nor muscle MRI or biopsy.^25^ Therefore, muscle involvement could be a common theme in spectrinopathy-related loss-of-function variants.

In conclusion, through an international effort we provide proof for *SPTAN1* loss-of-function variants leading to an early childhood onset distal weakness phenotype due to primary muscle involvement. Further research is needed to better understand the role of alpha-2-spectrin in skeletal muscle fibers and possible interactions with neuronal alpha-2-spectrin.

## Data Availability

The raw NGS data cannot be made publicly available for confidentiality policies in the consortia involved. All other data generated during this study is available upon reasonable request.

## ACKNOWLEDGEMENTS

The authors thank all patients for their participation in the study. The authors would like to acknowledge the contributions of Dr. Alan Beggs, Dr. Sarah Neuhaus, and Dr. A. Reghan Foley for their assistance with the evaluations of families H and K. The preliminary results and conclusions of this study was presented orally at the 5th Joint Meeting of the Belgian-Dutch Neuromuscular Study Club and the Reference Center for Neuromuscular Diseases in Vaals in 2024 titled “Heterozygous *SPTAN1* loss-of-function variants cause early childhood onset distal myopathy”. Presented orally and as a scientific poster at the World Muscle Society in Charleston in 2023 titled “Heterozygous *SPTAN1* frameshift mutations cause distal myopathy with neurogenic features”.^26^ Presented orally and as a scientific poster at the SOLVE-RD final meeting in Prague in 2023 titled “Heterozygous *SPTAN1* frameshift mutations cause distal myopathy with neurogenic features”. Abstracts of these presentations were published.

