## Supplemental material for "Heterozygous loss-of-function variants in SPTAN1 cause a novel early childhood onset distal myopathy with chronic neurogenic features"

**Supplementary methodology RNAseq N:II:1**

Total RNA was extracted from N:II:1 and control skeletal muscle biopsies (~15-50 mg) using the RNeasy Fibrous Tissue Mini Kit (Qiagen, Hilden, Germany) according to the manufacturer’s instructions. Strand-specific Poly-A+ RNA libraries were prepared from extracted RNA using the Agilent SureSelect XT library preparation kit (Agilent, Santa Clara, CA, USA). QC was performed using TapeStation 4200 (Agilent, CA, USA) and Qubit 4 Fluorometer (Thermo Fischer Scientific, Waltham, Massachusetts, USA), as well as QC sequencing on the Illumina iSeq 100 flowcell (Illumina, San Diego, CA, USA). These strand-specific libraries were sequenced on Illumina NovaSeq 6000 to produce paired-end 150 bp reads and an average of 50 million read pairs per sample. Adaptor sequences were removed and demultiplexed FASTQ files were provided by Genomics WA (Western Australia) for download and further analysis. FASTQ files were processed, including read quality control and alignment, using the nf-core/rnaseq pipeline ([https://nf-co.re/rnaseq](https://nf-co.re/rnaseq/1.3)), v3.8.1. Trimmed reads were aligned to the NCBI GRCh38 human reference genome using STAR v2.7.10a^1^ (STAR, RRID: SCR_004463). We used DROP v1.0.3 ^2^, as previously described ^3^ to analyse aberrant gene expression amongst a cohort of 150 skeletal muscle RNA-seq from rare muscle disease patients and unaffected controls. DROP leverages OUTRIDER ^4^, which uses a denoising autoencoder to control co-variation before fitting each gene over all samples via negative binomial distribution. Multiple testing correction was done across all genes per sample using DROP’s in-built Benjamini-Yekutieli’s false discovery rate (FDR) method. Plots were prepared using R (v4.1.3) in RStudio. The splicing pattern and expression of *SPTAN1* in N:II:1 was visualised using ggsashimi ^5^.
